# The common H202D variant in GDF-15 does not affect its bioactivity but can significantly interfere with measurement of its circulating levels

**DOI:** 10.1101/2022.01.03.22268655

**Authors:** Y Karusheva, M Ratcliff, A Melvin, A Mörseburg, N Sattar, P Barker, K Burling, A Backmark, R Roth, L Jermutus, E Guiu-Jurado, M Blüher, P Welsh, M Hyvönen, S O’Rahilly

## Abstract

Genetic variants in proteins can interfere with measurement of their circulating concentrations. Given the growing biomedical importance of GDF-15, we wished to establish whether a common histidine to aspartate variant present in position 6 of the mature GDF-15 protein (H202D variant) interfered with its measurement by two commonly used immunoassays. We first examined the detectability of recombinant monomers, homodimers and heterodimers of GDF-15 by assays and/or reagents used in two widely used immunoassays (Roche Elecsys® GDF-15 and the R&D antibody combinations used in their Quantikine® and DuoSet® ELISA’s). The Roche assay detected the H and D containing peptides similarly but the assays based on the R&D reagents consistently underreported concentrations of the D-containing variant peptide. Measurements of plasma concentrations of GDF-15 in genotyped human participants showed that the R&D reagents reported values in heterozygotes were ∼25% lower, and in homozygotes, 50% lower than the Roche assay. We finally studied the activation of the GDF-15 receptor, GFRAL-Ret, in a cell based assay and found that the activities of the HH and DD containing GDF-15 peptide were indistinguishable. These results have implications for the interpretation of genetic epidemiological studies which have used the R&D reagents to measure GDF-15, and for the emerging clinical use of GDF-15 as a diagnostic and prognostic biomarker. We provide correction equations, which may be of utility for the analysis of data generated with the R&D reagents where the genotype of the participants is known.

## Background

Growth-differentiation factor 15 (GDF-15) is a member of the transforming growth factor β (TGF-β) cytokine superfamily. Its circulating concentrations in man are elevated in a wide range of diseases such as cachexia ^1^, chemotherapy-related nausea ^2^ and vomiting, and hyperemesis gravidarum (HG) ^3^. It is also elevated in mitochondrial disorders ^4-6^, where it may have some diagnostic utility, and its levels are being investigated for potential prognostic value in some cancers and in heart failure ^7,8^. GDF-15 has recently gained further scientific and translational prominence with the discovery that its receptor is a GFRAL-RET heterodimer, which is found solely in the hindbrain ^9-12^. Activation of this receptor results in a reduction of food intake and body weight in several animal models ^13^. These findings have prompted exploration of GFRAL-Ret agonists as potential therapy for obesity and antagonists of GDF-15 or GFRAL in various forms of cachexia. While studies in rodents and primates have consistently demonstrated the weight loss effects of GDF-15, a recent Mendelian randomisation study did not support the idea that increased levels of GDF-15 (measured by immunoassay using the R&D antibodies^14^) reduced human BMI. Conversely, it was suggested that genetically determined increases in circulating GDF-15 might actually predispose to obesity. As there is a common coding variant in GDF-15, which changes residue 202 from histidine to aspartic acid ^15,16^ and is present in all human populations studied to date with a minor allele frequency ranging from 14% to 35% ^17^, we speculated that the discrepancy between pharmacological and Mendelian randomisation studies might be due to systematic bias in the measurement of GDF-15 by certain immunoassays. To test that hypothesis, we first examined the ability of the antibody combinations from two widely used assays for human GDF-15 to detect the H and D isoforms, using the relevant synthetic homodimers and heterodimers. Secondly, we studied human plasma samples from participants of known genotype. Finally, we undertook studies comparing the functional properties of the H and D forms of GDF-15 as activators of signalling through GFRAL-Ret.

## Methods

### Study participants

Circulating GDF-15 concentrations were measured in plasma from 173 participants (126 women, 47 men with BMI range: 27.5-70 kg/m^2^, age range: 21-74 years) from the Leipzig Obesity Biobank whose genotype at rs1058587 in the GDF-15 gene had previously been established from RNA sequencing data using adipose tissue obtained at surgery. Participants were randomly selected from the Biobank to represent an equal distribution of the genotype at rs1058587 ^18^. Plasma samples were collected in the context of elective laparoscopic abdominal surgeries as described previously ^19^. After extraction, plasma was immediately frozen in liquid nitrogen and stored at -80 °C. The study was approved by the Ethics Committee of the University of Leipzig (approval no: 159-12-21052012), and performed in accordance to the declaration of Helsinki. All subjects gave written informed consent before taking part in this study. Measurement of body composition and metabolic parameters was performed as described previously ^18,19^.

### Protein production

DNA encoding for mature GDF-15 was cloned into pBAT4 vector using *Nco*I and *Hind*III restriction sites, with the addition of a codon for an initiation methionine. The H202D mutation (histidine to aspartate variant present in position 6 of the mature GDF-15 protein) was introduced by PCR mutagenesis of the wild type construct. The mature GDF-15 proteins were expressed in *E. coli* in 2xYT medium at 37 °C, with all the protein forming insoluble inclusion bodies. The bacterial cell cultures were pelleted by centrifugation. Pelleted cells were resuspended in 30 ml lysis buffer (50 mM TRIS HCl pH 8.0, 5 mM EDTA with 10 mM dithiothreitol (DTT)) per 1 L of cell culture. Cell lysis was achieved under high pressure using an Emulsiflex C5 homogenizer, with 0.5% Triton-X added to the sample during lysis. The lysate was then incubated at room temperature with 0.2 mg of DNase and 4 mM MgCl_2_ for 30 minutes. The lysate was centrifuged at 15,000 g for 20 minutes and the supernatant discarded. Centrifugation and removal of the supernatant was repeated after resuspension with the following wash buffers; Buffer A (50 mM Tris pH 8.0, 5 mM EDTA, 10 mM DTT, 0.5% v/v Triton-X 100), Buffer A with 2 M NaCl and finally Buffer A without Triton-X 100. The purified inclusion body was pelleted by centrifugation and stored at -20 °C. Inclusion body pellets (from 1 L of bacterial cell culture) were re-suspended in 5 to 10 ml in 100 mM TCEP pH 7.2, then solubilised with the addition of 15 to 30 ml 8 M guanidine hydrochloride, 50 mM Tris-HCl pH 8.0 and 10 mM EDTA. Samples were then incubated at room temperature for 20 minutes, clarified by centrifugation and buffer exchanged into deionised 6 M urea with 20 mM HCl using Sephadex G25 column (Cytiva). The protein concentration was adjusted to ≤ 1 mg/ml by rapid dilution in urea buffer (100 mM TRIS pH 8.0, 1.2 M urea, 1 M PPS, 0.2 mM L-cystine) For refolding of the heterodimeric GDF-15 (HD), equal quantities of denatured H and D forms of GDF-15 were mixed before dilution in refolding buffer. This resulted in a “heterozygous mix” containing GDF-15 HD, HH and DD as confirmed by mass spectrometry (MS) with an approximate 1:2:1 ratio of HH:HD:DD.

Refolded GDF-15 was captured by ion-exchange chromatography using SOURCE 30S column (Cytiva) and fractions containing the disulfide linked dimer polished using reversed phase chromatography using an ACE5 C8-300, size 4.6×250 (HiChrom). Eluted fractions were analysed by non-reducing SDS-PAGE. Fractions with pure GDF-15 dimer were pooled together, with the concentration determined by UV absorption at 280 nM using calculated absorption coefficient. Aliquots of 250 µg were vacuum dried and stored at -80°C.

For subsequent assays, the proteins were suspended to 1 mg/ml in 10 mM HCl and diluted to desired concentration in buffer containing bovine serum albumin.

### Immunoassays

#### Roche Elecsys® GDF-15

Samples were analysed using the Roche Elecsys® e411 GDF-15 Cobas electrochemiluminescent immunoassay (Roche Diagnostics, Burgess Hill, UK). In the first incubation, GDF-15 in the sample, the biotinylated monoclonal GDF-□5Lspecific antibody and a second monoclonal GDF-15Lspecific antibody conjugated to rheuthinium form a sandwich complex. In the second incubation, after addition of streptavidin-coated microparticles, the complex becomes bound to the microparticles via interaction of biotin and streptavidin. The reaction mixture is aspirated into the measuring cell where the complex-microparticles are magnetically captured onto the surface of the electrode. Unbound substances are then removed with wash solution (ProCell II M). Application of a voltage to the electrode then induces electrochemiluminescent emission which is measured by a photomultiplier. Results are determined via a calibration curve which is instrument specifically generated by 2□point calibration and a master curve provided via the Cobas link. The limit of detection for the assay was 400 pg/ml, with an upper measurement limit of 20,000 pg/ml. Samples that were initially above the measuring range were diluted in Roche ‘Multiassay diluent’. Assays were calibrated and quality controlled using the manufacturer’s reagents. Coefficient of variation was 3.8% for the low control (at 1556 pg/ml) and 3.4% for the high control (at 7804 pg/ml)

#### R&D Quantikine® ELISA Assay

This is a quantitative sandwich enzyme immunoassay with a monoclonal antibody specific for GDF-15 pre-coated onto a microplate. Standards and samples are pipetted, in duplicate, into the wells and any GDF-15 present is bound by the immobilized antibody. After washing, a horse radish peroxidase (HRP) conjugated polyclonal antibody specific for GDF-15 is added to the wells. Following a wash to remove any unbound conjugate, a substrate solution is added to the wells and colour develops in proportion to the amount of GDF-15 bound in the initial step. The colour development is stopped with acid and the absorbance of the solution is measured on the Perkin Elmer Victor3 plate reader. The assay was controlled using in-house generated controls based on anonymised serum and plasma pools as used for the MSD R&D DuoSet® in-house assay.

The limit of detection of the assay is 17.6 pg/ml with an upper limit of the standard curve of 1500 pg/ml (manufacturer’s data). The between batch imprecision is 6.0% at 225 pg/ml, 4.7% at 442 pg/ml, and 5.6% at 900 pg/ml (manufacturer’s data).

#### MesoScale Discovery (MSD) R&D DuoSet® in-house assay

The assay is an in-house microtitre plate-based two-site electrochemiluminescence immunoassay using the MesoScale Discovery assay platform (MSD, Rockville, Maryland, USA). Antibodies and standards were purchased as a DuoSet® (catalogue number DY957) from R&D Systems (BioTechne, Abingdon, UK). Standard-bind MSD microtitre plates were coated overnight with monoclonal anti-GDF-15 capture antibody diluted in PBS. After coating, the plate was washed three times with PBS/Tween using a ThermoFisher automated plate washer and then blocked with MSD Blocker A. After blocking, 40 µl assay diluent plus 10 µl Standards, QCs and undiluted sample were added to the plate in duplicate. After 2 h incubation at room temperature on a plate shaker, the plate was washed three times with PBS/Tween. Biotinylated polyclonal goat anti-human GDF-15 detection antibody was diluted in MSD Diluent 100 and added to the plate. After 1 h incubation at room temperature on a plate shaker and washing with PBS/Tween, Sulpho-TAG labelled Streptavidin (MSD) diluted in MSD Diluent 100 was added to the plate. The plate was the incubated on a plate shaker for a further 30 minutes.

After another wash step, MSD read-buffer was added to all the wells and the plate was immediately read on the MSD s600 plate reader. Luminescence intensities for the standards were used to generate a standard curve using MSD’s Workbench software package. The assay was controlled using in-house generated controls based on anonymised serum and plasma pools.

The limit of detection of the assay is 3.0 pg/ml with an upper limit of the standard curve of 32,000 pg/ml (in-house data). The between batch imprecision is 9.8% at 352 pg/ml, 7.8% at 1476 pg/ml, 8.0% at 6593 pg/ml and 6.1% at 12134 pg/ml (in-house data).

#### GDF-15 Bioassay of Recombinant GDF-15 Protein Constructs

GDF-15 bioactivity assays were performed in HEK293S-SRF-RET/GFRAL cells. This stable cell line was modified to express full length human RET and GFRAL receptors as well as serum response factor-responsive luciferase reporter gene by AstraZeneca. Cells were grown and seeded in T75 flasks in DMEM GlutaMAX (Gibco) supplemented with 10% foetal bovine serum (FBS) (Gibco) and incubated for 30 h at 37°C, with 5% carbon dioxide. Post incubation, media was switched to serum free DMEM and cells were then incubated overnight under the same conditions. The following day, cells were detached using Accutase enzyme and then neutralised in DMEM with 1% FBS. Cells were then pelleted by centrifugation and resuspended in serum free DMEM with an additional 25 mM HEPES pH 7.2 (GDF-15 bioassay media). Cells in suspension were counted using a Cedex automatic cell counter (Roche) and Trypan Blue (Sigma) staining as standard. After counting, cells were then diluted to 500,000 cells/ml in GDF-15 bioassay media before plating in 384-well plates (15 µl/well). After plating, 5 µl of recombinant GDF-15 in GDF-15 bioassay media was then added to the cells at 4x the final concentration. Cells with recombinant protein samples were then incubated for 5 h at 37°C, with 5% carbon dioxide. After 5 h, the plate was then equilibrated to room temperature before the addition of Steady-Glo Luciferase Assay System solution (Promega) at 12µl/well and incubated for a further 20 minutes. The luminescence was measured using an EnVision plate reader (Perkin Elmer) with crosstalk correction applied. CHO derived recombinant Human GDF-15/MIC-1 (PeproTech 120-28C) was used as a positive control. Negative controls of protein sample buffers only were also screened to confirm absence of signalling activity. Triplicates of each dose were performed, and data was analysed in GraphPad 7.0 (Prism).

#### Regression equations

Passing-Bablok regressions ^20^ were run using the R package mcr (v.1.2.1) ^21^ for N = 173 patients to obtain correction equations between the MSD R&D DuoSet® in-house and Roche immunoassays, stratified by genotype.

## Results

### Recovery of the D variant in synthetic peptides

We synthesised three forms of GDF-15 (HH homodimers, DD homodimers and HD heterodimers, the result of a mixture of HH:HD:DD in ratio 1:2:1). Equal amounts of each dimer were serially diluted and assayed using the Roche Elecsys® and the R&D Quantikine® ELISA and MSD R&D DuoSet® immunoassays (see Methods). Table 1 shows the data for the sample with a theoretical concentration of 667 pg/ml. This is the only concentration which was within the working ranges of all three assays. The full data set is included in the Suppl. Table 1. Recoveries of HH and HD with the Roche Elecsys® assay were very similar with a slight reduction (8-15%) in recovery when assaying the D dimer (Table 1 + Suppl. Table 1). With the Roche Elecsys® assay reported results were proportional to dilution from 2000 to 667 pg/ml. The Roche Elecsys® assay has a lower limit of detection of 400 pg/ml and therefore further dilution were reported as <400 pg/ml as expected (Suppl. Table 1). With the MSD R&D DuoSet® immunoassays, which has a lower limit of detection of 3.0 pg/ml the measurements were also proportionate to the dilutions, the percentage recovery was similar across all dilutions. However, there was a marked difference in measured levels between the types of dimer (Table 1 and Suppl. Table 1).

**Table 1.**
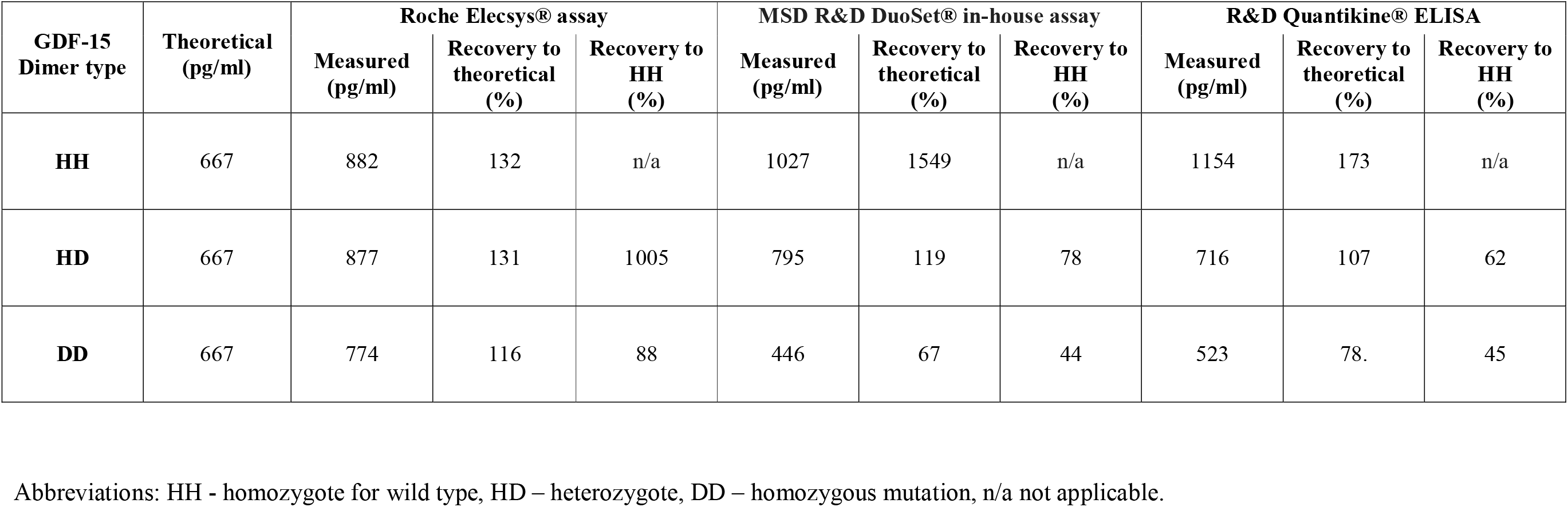
Percentage recovery of synthetic GDF-15 dimers HH, HD and DD using three immunoassays.

| **GDF-15**<br>**Dimer type** | **Theoretical**<br>**(pg/ml)** | **Roche Elecsys® assay** |  |  | **MSD R&D DuoSet® in-house assay** |  |  | **R&D Quantikine® ELISA** |  |  |
| --- | --- | --- | --- | --- | --- | --- | --- | --- | --- | --- |
|  |  | **Measured**<br>**(pg/ml)** | **Recovery to**<br>**theoretical**<br>**(%)** | **Recovery to**<br>**HH**<br>**(%)** | **Measured**<br>**(pg/ml)** | **Recovery to**<br>**theoretical**<br>**(%)** | **Recovery to**<br>**HH**<br>**(%)** | **Measured**<br>**(pg/ml)** | **Recovery to**<br>**theoretical**<br>**(%)** | **Recovery to**<br>**HH**<br>**(%)** |
| **HH** | 667 | 882 | 132 | n/a | 1027 | 1549 | n/a | 1154 | 173 | n/a |
| **HD** | 667 | 877 | 131 | 1005 | 795 | 119 | 78 | 716 | 107 | 62 |
| **DD** | 667 | 774 | 116 | 88 | 446 | 67 | 44 | 523 | 78. | 45 |

Abbreviations: HH - homozygote for wild type, HD – heterozygote, DD – homozygous mutation, n/a not applicable.

The R&D Quantikine® ELISA and MSD R&D DuoSet® immunoassays gave comparable results. Recovery of DD dimers with the both of these assays was approximately 45% of that seen with HH dimers, and between 62 and 78% with the HD dimers (Table 1 and Suppl. Table 1).

### Recovery of the D variant in human plasma samples

We compared the circulating concentrations of GDF-15 as measured by the Roche Elecsys® and MSD R&D DuoSet® immunoassays in plasma samples from 173 participants (126 women, 47 men) from the Leipzig Obesity Biobank whose genotype at rs1058587 in the *GDF-15* gene had previously been established from RNA sequencing data using adipose tissue obtained at surgery ^18^. We selected samples from 61 HH homozygotes, 59 HD heterozygotes and 53 DD homozygotes.

Fig. 1 shows the results of the Mean +-SEM of plasma GDF-15 in participants with all three genotypes. This was consistent with our findings with the synthetic peptides. The concentrations of GDF-15 reported using MSD R&D DuoSet® immunoassay were ∼9% lower in HH, 32% lower in HD and 62% lower in DD than those measured by the Roche Elecsys® assay.

**Figure 1.**
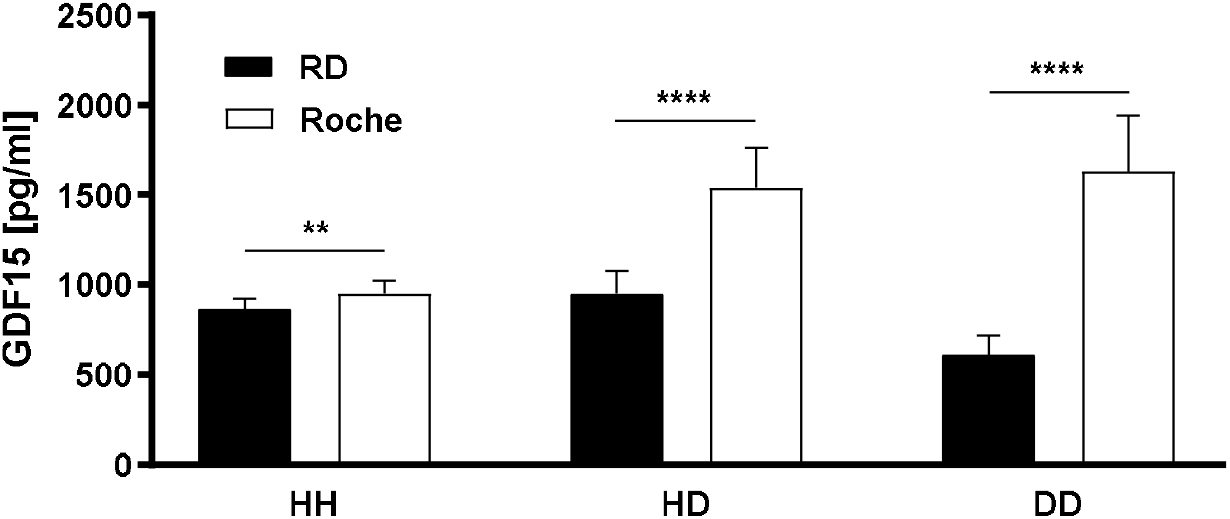
Circulating levels of GDF-15 measured in genotyped human participants using MSD R&D DuoSet® and Roche Elecsys® immunoassays. Serum GDF-15 concentrations in genotyped 173 participants of the Leipzig Obesity Biobank measured by MSD R&D DuoSet**®** immunoassay (RD) against serum GDF-15 measured by the Roche Elecsys® immunoassay (Roche) in the same individuals. Abbreviations: Wild type (HH) n=61, heterozygote (HD) n=59, homozygote (DD) n=53. Data are shown as mean ± SEM. \*\**P*<0.01, \*\*\*\**P*<0.0001.

From the data produced using the synthetic peptide it appears that the Roche Elecsys® assay is able to detect the three forms of GDF-15 approximately equally. From this we can then attempt to establish a correction factor that could be used, in situations where genotype is known, to convert MSD R&D DuoSet® immunoassay results to an estimation of the Roche Elecsys® assay for samples from individuals who are homozygous or heterozygous for the D mutation. Figure 2 provides a graphical representation the relationship between GDF-15 levels measured using the two assays in the same individuals. Passing Bablok (PB) regression ^20^ was applied to obtain correction equations as it is more resistant to outliers than simple linear regression. The results of the PB regression (Figure 2) were consistent with the outcomes of the t-tests (Figure 1). Equations 1 and 2 describe the necessary corrections for HD heterozygotes and HH homozygotes respectively, confidence intervals for the regression parameters are given in Figure 2. The 95% confidence intervals for the slopes do not overlap, consistent with strong evidence against the null hypothesis that the slopes are the same gradient.

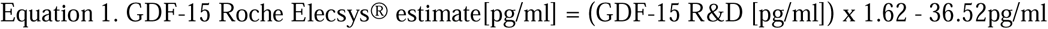

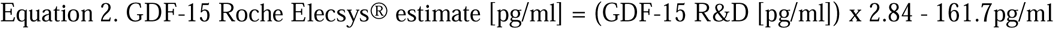

**Figure 2.**
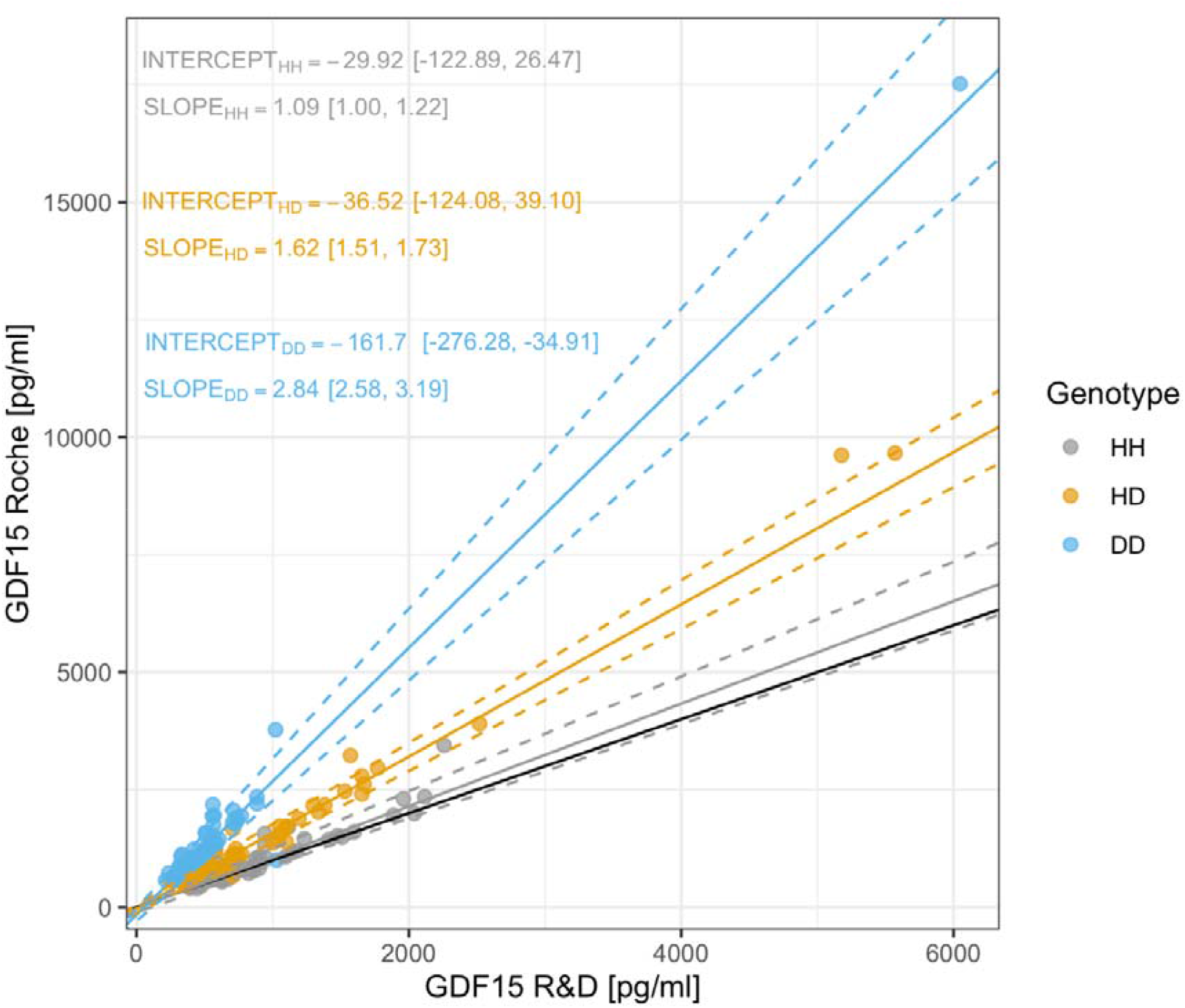
Serum GDF-15 concentrations in genotyped persons (n = 173) measured by MSD R&D DuoSet® immunoassay plotted against serum GDF-15 measured by Roche Elecsys® immunoassay in the same individual. Passing Bablok regression lines are given, line colours represent the genotype of the respective person at rs1058587 and dotted lines the 95% CIs. Numerical values for intercepts and slopes with 95% CIs in square brackets are given. The solid black line denotes the diagonal. Abbreviations: HH - homozygote for wild type, HD – heterozygote, DD – homozygous mutation.

### H and D containing forms of GDF-15 have equivalent bioactivity on the GFRAL-Ret receptor

HEK293S cells stably expressing both components of the GDF-15 Receptor, GFRAL and Ret, and containing a serum response factor responsive luciferase reporter gene were exposed to varying concentrations of synthetic forms of GDF-15 to establish their bioactivity. Both HH and DD homodimers of GDF-15 showed indistinguishable activity in their dose-response characteristics with EC_50_ values of 0.39 nM (95% CI 0.44-0.45 nM) for HH variant and 0.45 nM (95% CI 0.38-0.52 nM) for DD variant (Figure 3).

**Figure 3.**
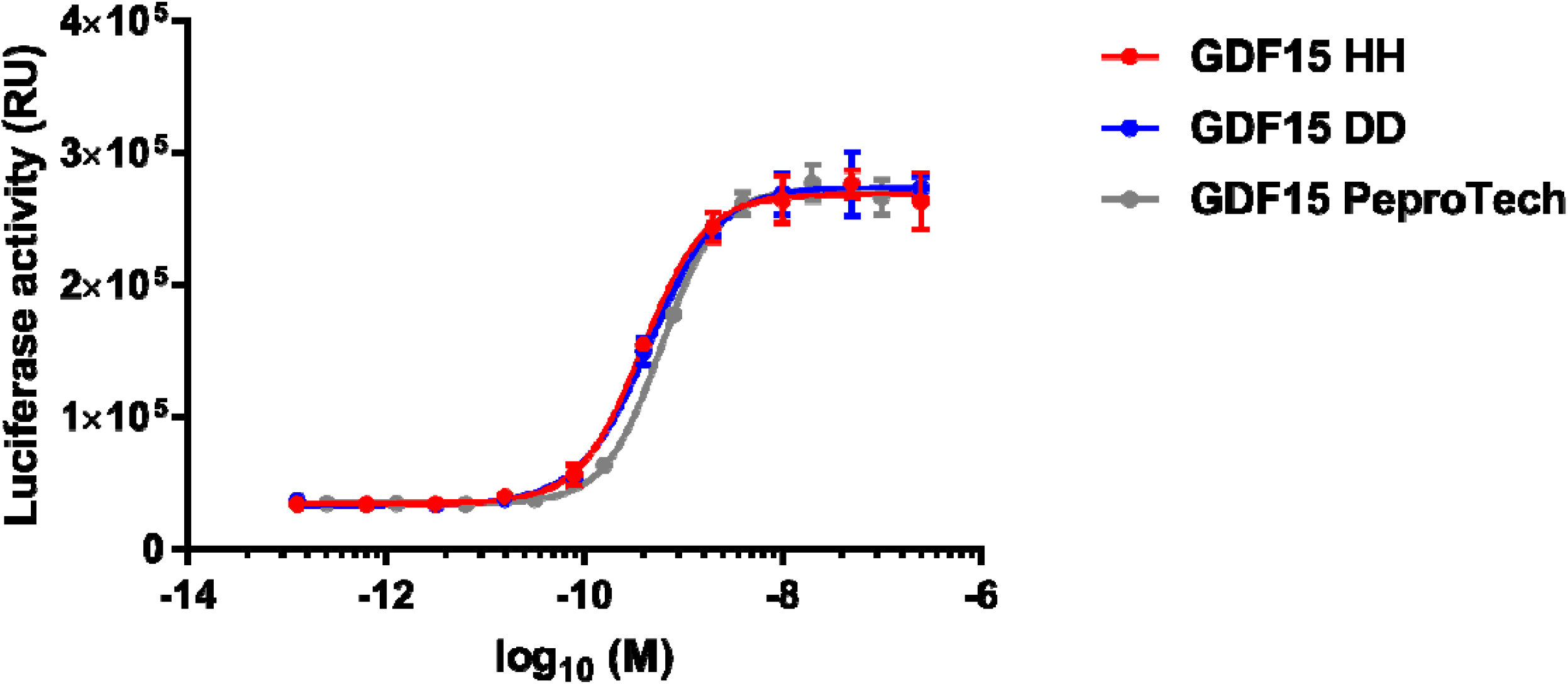
Bioactivity of H and D containing GDF-15 dimers on GFRAL-Ret signalling. Bioactivity of HH and DD variant mature GDF-15 in HEK293S-SRF-RET/GFRAL cells with luciferase reporter under serum responsive factor control. CHO derived recombinant human GDF-15/MIC-1 (PeproTech) was used as a positive control.

## Discussion

We have found that the reagents used in one frequently used immunoassay for GDF-15, from R&D Systems, systematically and seriously underestimate circulating concentrations of GDF-15 in people who carry one or two copies of the D allele at position 6 of the mature peptide (position 202 of the pro-peptide). In contrast, the Roche Elecsys® assay appears to be far less affected, though studies with *in vitro* peptides suggest that homozygote DD dimers may be under-recovered by around 10%. As the epitopes targeted by commercial immunoassays are proprietary information, we are not in a position to report whether this difference in assay performance reflects the selection of the antibodies used. However, residue 6 of the mature peptide is in a flexible and accessible region of the GDF-15 molecule and is likely to be a site against which antibodies are readily generated. This common amino acid polymorphism might also affect the accurate measurement of GDF-15 by other types of assay components, such as aptamers ^22^. A substantial proportion of the published literature regarding circulating levels of GDF-15, including work from our own laboratory, has used methodology based on the R&D antibodies and the conclusions of such studies will need to be re-evaluated on the basis of this new information. In general, the assay appears to perform well within a given individual, so findings that measure levels in research participants before and after any experimental manipulations or serially over time should retain their validity. However, any cross sectional studies comparing individuals will need to be viewed with caution as there is a serious risk that an inadvertent and unrecognised imbalance of genotypes between groups would be a critical source of inaccuracy.

Furthermore, clinical trials of GDF-15 antagonism are currently being undertaken in states of cachexia^23^ with entry criteria for the trials including elevated concentrations of GDF-15. The GDF-15 concentrations may be significantly underestimated in individuals with the D genotype if a genotype affected assay is used, resulting in the unnecessary exclusion of patients from the trial.

Genetic variants in and around the GDF-15 gene, one of which is the rs1058587 encoding the H202D variant are associated with the pregnancy-specific condition HG at genome wide levels of significance^24^. As GDF-15 administration in animals such as shrews and primates results in vomiting, it might have been expected that the SNPs would affect HG risk through increasing circulating levels. However, studies of circulating levels in relation to genotype have not always been directionally consistent with such an effect. The knowledge that the D variant has such a significant effect on measurement by certain assays formats provides a possible explanation for the puzzling findings reported to date. Similarly, when Karhunen et al ^14^ performed Mendelian randomisation studies asking whether genetic variants increasing GDF-15 levels were associated with BMI, their surprising conclusion that GDF-15 might actually increase body weight could again be explained by the fact that the assay used to measure GDF-15 used R&D reagents. Although we cannot definitely report that these assay inconsistencies explain inconsistencies in the published epidemiological data, we provide a correction factor which we propose can be usefully employed to re-analyse data from genotyped individuals in such Mendelian randomisation studies. It is very likely that the H202D variant under recovery will also occur using other assay methodologies that have been employed in large-scale proteomic studies ^21^.

A further level of complexity would be provided if there were differences in bioactivity between the H and D variants. The known structure of the GDF-15-GFRAL complex indicates that position 6 in the mature growth factor is not critically involved in this interaction as the data presented here support the idea that they are bioequivalent. It is at least theoretically possible that the H and D variants could act in subtly different ways *in vivo*, such as through differential interaction with components of the extracellular matrix.

In conclusion, the genetic variant of GDF-15, present in 15-30% of people worldwide, substantially affects its measurement by one or more widely used assays with implications for the interpretation of studies employing such assays.

## Data Availability

All data produced in the present study are available upon reasonable request to the authors.

