## Supplemental Table 1 for "The common H202D variant in GDF-15 does not affect its bioactivity but can significantly interfere with measurement of its circulating levels"

**Supplementary Table 1. Percentage recovery of dilutions of synthetic GDF-15 dimers HH, HD and DD using three immunoassays.**

| **GDF-15 Dimmer type** | **Theoretical (pg/ml)** | **Roche Elecsys® assay** | | | **MSD R&D DuoSet® in-house assay** | | | **R&D Quantikine® ELISA** | | |
| --- | --- | --- | --- | --- | --- | --- | --- | --- | --- | --- |
|  |  | **Measured**  **(pg/ml)** | **Recovery to theoretical**  **(%)** | **Recovery to HH**  **(%)** | **Measured**  **(pg/ml)** | **Recovery to theoretical**  **(%)** | **Recovery to HH**  **(%)** | **Measured**  **(pg/ml)** | **Recovery to theoretical**  **(%)** | **Recovery to HH**  **(%)** |
| **HH** | 2000 | 2721 | 136 | n/a | 3329 | 166 | n/a | >1500 | n/a | n/a |
|  | 667 | 882 | 132 | n/a | 1027 | 154 | n/a | 1154 | 173 | n/a |
|  | 222 | <400 | n/a | n/a | 367 | 165 | n/a | 363 | 164 | n/a |
|  | 74 | <400 | n/a | n/a | 121 | 163 | n/a | 120 | 162 | n/a |
| **HD** | 2000 | 2773 | 139 | 102 | 2384 | 119 | 72 | >1500 | n/a | n/a |
|  | 667 | 87 | 132 | 100 | 795 | 119 | 77 | 716 | 107 | 62 |
|  | 222 | <400 | n/a | n/a | 282 | 127 | 77 | 212 | 96 | 58 |
|  | 74 | <400 | n/a | n/a | 88 | 118 | 73 | 75 | 101 | 63 |
| **DD** | 2000 | 2327 | 116 | 84 | 1269 | 63 | 53 | >1500 | n/a | n/a |
|  | 667 | 774 | 116 | 88 | 446 | 67 | 56 | 523 | 78 | 73 |
|  | 222 | <400 | n/a | n/a | 155 | 70 | 55 | 176 | 79 | 83 |
|  | 74 | <400 | n/a | n/a | 55 | 74 | 62 | 65 | 88 | 87 |

Abbreviations: HH - homozygote for wild type, HD – heterozygote, DD – homozygous mutation, n/a not applicable.
